# Proteomics analysis of plasma for risk of sepsis: Findings from the Atherosclerosis Risk in Communities Study

**DOI:** 10.1101/2025.03.07.25323594

**Authors:** Junichi Ishigami, Xiao Hu, Pascal Schlosser, Thomas R Austin, Jingsha Chen, Bruce M. Psaty, David Dowdy, Christie Ballantyne, Morgan Grams, Josef Coresh, James S Floyd, Kunihiro Matsushita

## Abstract

**Objective:** Sepsis is a serious condition resulting from infection associated with high mortality. A high-throughput analysis of circulating blood proteins may provide mechanistic insight and potent therapeutic targets for the prevention of sepsis.

**Patients and Methods:** We used multivariable Cox regression analysis to examine the association of 4,955 plasma proteins measured by SomaScan with the risk of incident sepsis, defined by hospital discharge with a primary diagnosis code for sepsis, among 11,065 participants of the Atherosclerosis Risk in Communities (ARIC) Study (visit 3 in 1993-95; mean age, 60.1 years, 54.4% female, 21.0% Black). Proteins (false discovery rate [FDR] of P <0.05) discovered at visit 3 were replicated using data at visit 5 (n=4,869 in 2011-13: mean age, 75.5 years) and in the Cardiovascular Health Study (CHS) (n=3,512 in 1992-93; mean age, 74.5 years). Canonical pathways were identified by enrichment analyses.

**Results:** We identified 669 proteins associated with the risk of sepsis in the ARIC visit 3 cohort. Of those, 175 proteins were significantly associated with sepsis in the visit 5 cohort. Of the 175 proteins, 90 proteins were replicated in an external replication cohort of CHS. The top 20 proteins ranked by P value were relevant to acute inflammatory signaling in innate immunity (e.g., GDF15, EGFR, CNTN1, HDGF, NBL1, TNFRSF1A, TFRSF1B, IL15RA, SLAMF1). Pathway analyses implicated activation of pro-inflammatory pathways (e.g., cytokine storm signaling) as well as inhibition of anti-inflammatory pathways (e.g., Liver X Receptor/Retinoid X Receptor [LXR/RXR] Activation), which also play relevant roles in lipid metabolism.

**Conclusions:** In this large-scale proteomics analysis, levels of acute inflammatory proteins measured during routine visits were associated with the subsequent incidence of sepsis. An increased risk of sepsis associated with the inhibition of anti-inflammatory pathways, such as LXR/RXR Activation warrants further mechanistic investigation.

## Introduction

Sepsis is a serious public health concern associated with morbidity and mortality. More than 1.7 million adults develop septicemia in the US each year,^1^ constituting the leading cause of death during hospitalization.^2^ Sepsis disproportionally affects immunocompromised hosts, such as older adults and individuals with chronic conditions (e.g., diabetes and chronic kidney disease).^3,4^ Altered metabolic changes in these conditions, such as the accumulation of glycated and uremic proteins in the bloodstream, may underlie an increased susceptibility to sepsis. Previous studies used a traditional targeted approach to associate selected blood biomarkers, such as interleukin-6 (IL-6) and tumor necrosis factor-α (TNFα), with the incidence of sepsis.^5–7^ However, these studies may provide limited mechanistic insight due to the complex pathophysiology of sepsis, which involves many proteins as well as multiple signaling pathways and transcription regulators.

A recent development of omics technology, a high-throughput analysis to relate thousands of proteins to clinical phenotypes, has enabled us to comprehensively interrogate proteomic signatures for a better understanding of disease pathophysiology. Here, we applied a proteomics study for the risk of sepsis, using data from the Atherosclerosis Risk in Communities (ARIC) Study. We first discovered plasma proteins associated with the risk of sepsis in ARIC at visit 3 (1993-95) and replicated the findings in ARIC visit 5 (2011-13) and the Cardiovascular Health Study (CHS) (1992-93). We then performed a pathway analysis to identify canonical pathways and upstream regulators relevant to risk of sepsis. We further performed a Mendelian randomization analysis to examine causal links between proteins and the risk of sepsis. Finally, we assessed the utility of proteome biomarkers to predict the risk of sepsis.

## Methods

### Study population

The Atherosclerosis Risk in Communities (ARIC) Study is a community-based cohort of US adults.^8,9^ Study visits occurred in 1987-1989 (visit 1),1990-1992 (visit 2), 1993-1995 (visit 3), 1996-1998 (visit 4), 2011-2013 (visit 5), and 2016-2017 (visit 6). After visit 6, participants were invited to visit study sites annually to biennially. In the ARIC Study, blood protein levels were measured at visit 3 and 5. For the primary analysis, we used data at visit 3 as baseline since this visit had a larger sample size and longer follow-up. Data at visit 5 were used as an internal and temporal replication cohort. At visit 3, we excluded those who were self-identified as other than Black or White race due to the small sample size (n=38); had missing covariates of interest (n=361), had history of sepsis prior to baseline visit (n=18) and missing information for incident sepsis (n=1). After additional exclusions of participants missing proteomics data (n=1404), the primary analytic cohort consisted of 11,065 middle-aged participants at visit 3 (mean age, 60.1 years, 54.4% female, 21.0% Black) (**Figure S1**). Applying the same exclusion criteria, the internal replication cohort consisted of 4,869 older-age participants at visit 5 (mean age, 75.5 years, 56.8% female, 18.8% Black) (**Figure S1**).

We used data from the Cardiovascular Health Study (CHS) to externally replicate our findings in ARIC. The CHS is a population-based longitudinal study of coronary heart disease and stroke in adults aged 65 years and older. At baseline (1989-90), 5201 individuals were enrolled. An additional 687 African Americans were recruited during 1992-93. Proteome measurements were obtained from blood plasma samples collected from 3,678 participants during the 1992-93 exam.^10^

For both the ARIC Study and the CHS, written informed consent was obtained from all participants, and the institutional review board at each study site approved the study.

### Proteomics

In ARIC, visit 3 and 5 plasma samples were collected according to study protocol and were stored at −80°C until they were analyzed in 2018-2019. Plasma proteins were measured using a multiplexed modified DNA-based aptamer technology version 4 platform (SomaScan version 4; SomaLogic, Boulder, CO).^11,12^ Slow off-rate modified aptamer (SOMAmer) reagents captured proteins from blood samples, then the SOMAmer reagents were measured in fluorescent arrays. The relative concentration of proteins was derived from the concentration of SOMAmer reagents. Two-step quality control processes were performed first by SomaLogic followed by the ARIC investigators, as detailed in the previous report.^13^ Of 5,284 SOMA-aptamers, 4,955 passed quality control and were used in the current analysis.

In CHS, fasting blood samples were collected according to study protocol and were stored at −70°C until they were analyzed. Previously unthawed EDTA-plasma samples were used for proteomic profiling with the SomaScan version 4 panel in 3188 participants in CHS, consisting of 5284 aptamers In an additional 490 participants, proteomic profiling was performed with the SomaScan version 4.1 panel consisting of 7596 aptamers. Scaling factors provided by SomaLogic were applied to aptamers in the version 4.1 panel that overlap with those in the version 4.0 panel allowing for jointly analyzing the data. Aptamers marked as “deprecated” (indicating a retired aptamer) or “non-human,” and fagged for poor quality were excluded from the analysis.

### Outcome

Our primary outcome was incident hospitalization with sepsis (hereafter “incident sepsis”), which was captured through the International Classification of Diseases, Ninth or Tenth Revision, Clinical Modification (ICD-9/10-CM) on hospital discharge records (**Table S1**). In both the ARIC and CHS, research staff asked participants or their proxies through annual phone calls or during study site visits whether they had been hospitalized since the last contact.^14^ Active surveillance ascertained all available hospitalization records including the list of discharge diagnoses and corresponding ICD codes.

For the present study, we defined incident sepsis as recording of sepsis at the primary diagnostic position. This approach assumes that sepsis was the primary reason for hospitalization or primary clinical problem during the hospitalization. We captured concomitant ICD9/10 codes for infections such as pneumonia, urinary tract infections, gastrointestinal infections, and cellulitis and osteomyelitis (**Table S2**) and causative pathogens of sepsis when there were ICD-9/10 codes for pathogen-specific infections (e.g., Streptococcus pneumoniae) (**Table S3**). To avoid double-counting events between the analyses of visit 3 and visit 5, we applied specific criteria. If a participant attended both visit 3 and visit 5, we censored the data at the date of visit 5 for the primary analysis that used visit 3 as the baseline. For participants who attended visit 3 but did not attend visit 5, they were censored at the last date of visit 5.

Other censoring criteria included participants who had an event of interest, died, were lost to follow-up, or were administratively censored on December 31, 2019, whichever occurred first.

### Covariates

Age, sex, race, field center, education attainment and smoking status (never or ever) were self-reported. Education attainment were categorized as basic (less than completed high school), intermediate (high school or vocational school), or advanced (at least some colledge). Body mass index was calculated as body weight in kilograms divided by height in meters squared.

Diabetes was defined as fasting blood glucose ≥126 mg/dL, random blood glucose ≥200 mg/dL, self-reported physician diagnosis of diabetes, or use of antidiabetic medication. Hypertension was defined as systolic blood pressure ≥140 mmHg, diastolic blood pressure ≥90 mmHg, or use of antihypertensive medication within past two weeks. Estimated glomerular filtration rate (eGFR) was calculated with the CKD EPI creatinine equation.^15^ History of coronary heart disease, heart failure, and stroke were based on self-report, hospital discharge records, and physicians’ adjudication, as appropriate.^8^

### Statistical analysis

#### Protein levels and risk of incident sepsis

We used multivariable Cox proportional hazard models to estimate hazard ratios (HRs) and their 95% confidence intervals (CIs) for incident sepsis. All protein measurements were treated on a continuous scale and log base 2 transformed. The models were adjusted for age, sex, race, center, education attainment, body mass index, smoking status, eGFR, diabetes, hypertension, prevalent coronary heart disease, prevalent stroke, and prevalent heart failure. To address multiple comparisons, we adjusted P-values for significance using the Benjamin-Hochberg procedure with a false discovery rate (FDR) set at <5%. The analysis was first performed in the ARIC visit 3 cohort (i.e., discovery cohort), and proteins with significant associations were then evaluated in the ARIC visit 5 cohort (i.e., internal replication cohort). Proteins significantly associated with sepsis in the ARIC visit 5 cohort were subsequently replicated in the external CHS cohort.

### Pathway analysis

We used Ingenuity Pathway Analysis (IPA, QIAGEN Inc.) to examine the biological mechanisms involved in sepsis. IPA is a web-based application that contains a large, curated database of molecular interactions and gene-to-phenotype associations knowledge.^13^ For this analysis, we used the proteins that were significantly associated with sepsis both at ARIC visit 3 and 5. Pathway enrichment was assessed by P-value and z-score. P-value is based on right tail Fisher’s exact test. Threshold for statistical significance was a P value of <0.05 after Benjamini– Hochberg FDR adjustment. The threshold for Z-value was absolute z-scores of ≥2.^16^

### Causal inference analysis

To explore the causal role of proteins associated with risk of sepsis, we performed a protein-wide association study (PWAS). In brief, a PWAS is a Mendelian randomization analysis that utilizes genetic instruments to assess a possible causal effect of the protein on the risk of sepsis.^17,18^ In the ARIC Study, a previous study related protein levels to cis-protein quantitative trait loci (pQTL) and developed models to genetic variants in the encoding region (pQTL) and developed models to infer putatively causal effects of these proteins on other traits by Mendelian randomization.^19^ We utilized the FUSION workflow,^20^ incorporating elastic net modeling, with weights derived from the European ancestry subpopulation of the ARIC study. These weights were then combined with the corresponding European ancestry in-sample LD reference.^19^ PWAS approaches generally explain a greater amount of protein variation than single pQTLs alone and minimize horizontal pleiotropy due to reliance on cis-pQTLs only (and not trans-pQTLs). By combining PWAS model weights with summary statistics from large genome-wide association studies (GWAS) of select phenotypes, evidence of potential causal relationships can be evaluated. In the current study, summary statistics from a GWAS of the ICD-9 code of 038 (Sepsis) in UK Biobank were combined with protein models from ARIC.^21^ Statistical significance was determined using the Benjamin-Hochberg procedure with a FDR <0.05.

### Prediction model for sepsis risk

To examine the ability of proteins to predict the risk of sepsis, we developed models using ARIC visit 3 data to predict the 10-year risk of incident sepsis. The first (“base”) model included variables of age, sex, race, education attainment, body mass index, smoking status, eGFR, diabetes, hypertension, prevalent coronary heart disease, prevalent stroke, and prevalent heart failure. The second (“LASSO”) model included proteins that were selected based on LASSO regression analysis, in addition to covariates for the base model.

For each of the base and LASSO models, we ran Cox proportional analyses and calculated the Harrel’s c-statistics. In terms of the validation,^22^ we applied the same β-coefficients derived from the discovery cohort but using baseline hazard of the validation dataset (i.e., recalibration).

Then, we assessed whether the addition of the same set of proteins could improve prediction by refitting the prediction models in the validation cohorts. We evaluated the cumulative risk of sepsis, accounting for death as a competing risk using the Fine and Gray subdistribution hazard model to estimate the cumulative incidence function of sepsis over time, considering the competing risk of death. We generated the calibration plots to show 10-year predicted and observed risks. All statistical analyses were performed using Stata 18.0 (StataCorp, College Station, TX) and R statistical software (v4.1.1; R Core Team 2021).

## Results

### Proteins associated with sepsis

In ARIC visit 3 cohort (n=11,065, mean age, 60.1 [SD, 5.7], 54.4% female, 21.0% Black) (**Table S4**), there were 463 cases of incident sepsis prior to visit 5 during a median follow-up of 17.3 (IQI, 14.5 and 18.5) years. Crude incidence rate per 1,000 person-years was 2.7 (95%CI, 2.5 to 2.9). Causative organisms were documented in 29% of these cases, including 17% caused by gram negative rod and 11% caused by gram positive cocci (**Figure S2**). Along with sepsis, concomitant infections were documented in 69% of the cases, with the most common infection being pneumonia (34%), followed by urinary traction infections (25%), cellulitis and osteomyelitis (6%), and gastrointestinal tract infections (4%) (**Figure S3**).

In multivariable Cox analysis, 669 of 4,955 proteins were significantly associated with incident sepsis using FDR-corrected P value of <0.05 (“significant proteins”) (**Figure 1, Table S5, Table S6**). We then assessed the associations of these 669 proteins with sepsis in ARIC visit 5 cohort, which baseline occurred approximately 18 years after visit 3 (n= 4,869, mean age, 75.5 years, 56.8% female, 18.8% Black). During a median follow-up of 7.2 (IQI, 5.6 and 7.8) years after visit 5, there were 357 cases of incident sepsis: of the 669 proteins identified in the visit 3 cohort, 175 proteins remained significantly associated with sepsis in the visit 5 cohort (**Table S5, Table S7**). These 175 proteins were further evaluated in the external CHS cohort (n=3512, mean age 74.5, 60.8% female, 18.2% Black). In CHS, there were 321 cases of hospitalization with sepsis during a median follow-up of 12.5 (IQI, 7.7 and 18.2) years: Of the 175 proteins associated with sepsis in both ARIC visit 3 and ARIC visit 5, 90 proteins remained significantly associated with sepsis in CHS (**Figure 2** and **Table S7**).

**Figure 1:**
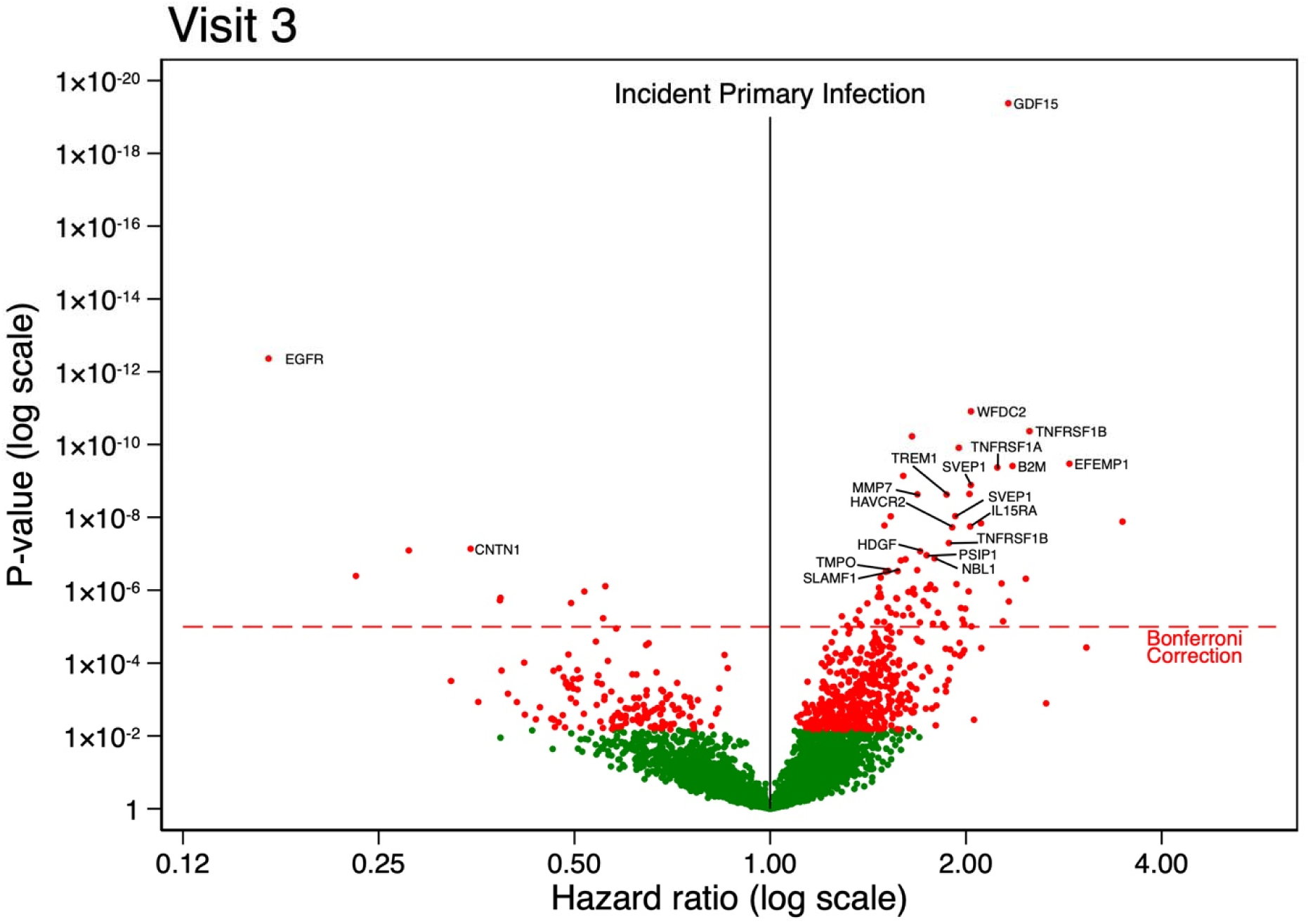
Volcano plots for risk of sepsis in ARIC visit 3 cohort. The models were adjusted for age, sex, race, center, education attainment, body mass index, smoking status, eGFR, diabetes, hypertension, prevalent coronary heart disease, prevalent stroke, and prevalent heart failure. Red dots indicate statistical significance by FDR criteria, green dots indicate non-significance by FDR criteria. Top 20 proteins ranked by P-values are labeled in the volcano plots.

**Figure 2:**
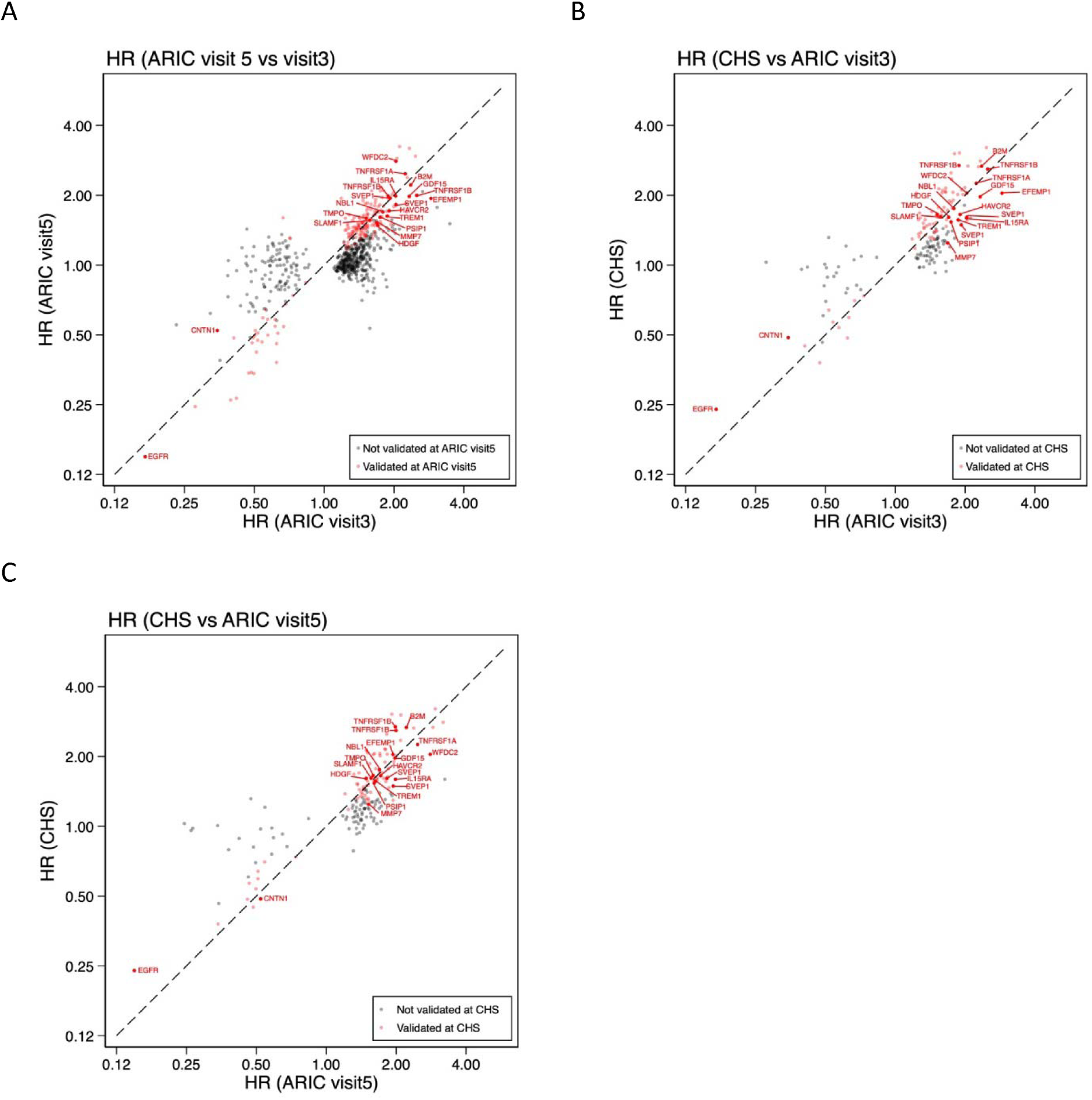
Scatter plots comparing adjusted hazard ratios for sepsis between primary cohort (ARIC visit 3) and replication cohort (CHS). The models were adjusted for age, sex, race, education attainment, body mass index, smoking status, eGFR, diabetes, hypertension, prevalent coronary heart disease, prevalent stroke, and prevalent heart failure.

Of these 90 proteins, the 20 with the lowest p-values at visit 3 included 18 unique proteins, as shown in **Table 1**. These proteins involved in innate immunity (i.e., initial response to pathogens), such as growth/proliferation factors (GDF15,^23^ EGFR,^24^ CNTN1,^25^ HDGF,^26^ NBL1^27^); cytokines or their receptors (TNFRSF1A/1B,^28^ IL15RA^29^, SLAMF1^30^); or immune modulators (WFDC2,^31^ B2M,^32^ MMP7,^33^ TREM1,^34^ HAVCR2,^35^ TMPO^36^). PSIP1 is an essential protein for human immunodeficiency virus (HIV) integration.^37^ EFEMP1^38^ SVEP1^39^ are extracellular matrix glycoproteins that interact with integrins to mediate immune cell communications. For the directionality of the association, EGFR and CNTN1 were associated with a reduced risk of sepsis while the rest of the 16 proteins were associated with an increased risk of sepsis.

**Table 1:** Top 20 proteins associated with sepsis risk in both ARIC and CHS cohorts. Due to duplicate proteins (SVEP1 and TNFRSF1B), there were 18 unique proteins. The models were adjusted for age, sex, race, education attainment, body mass index, smoking status, eGFR, diabetes, hypertension, prevalent coronary heart disease, prevalent stroke, and prevalent heart failure. Statistical significance was based on false discovery rate (FDR) P<0.05.

| Gene name | Full protein name | ARIC visit 3 |  |  | ARIC visit 5 |  |  | CHS |  |  |
| --- | --- | --- | --- | --- | --- | --- | --- | --- | --- | --- |
|  |  | HR | 95%CI | P-value | HR | 95%CI | P-value | HR | 95%CI | P-value |
| GDF15 | Growth/differentiation factor 15 | 2.32 | (1.94, 2.78) | 4.21E-20 | 1.98 | (1.61, 2.43) | 1.33E-10 | 1.97 | (1.48, 2.63) | 3.61E-06 |
| EGFR | Epidermal growth factor receptor | 0.17 | (0.10, 0.27) | 4.36E-13 | 0.15 | (0.09, 0.25) | 1.97E-13 | 0.24 | (0.12, 0.47) | 2.62E-05 |
| WFDC2 | WAP four-disulfide core domain protein 2 | 2.04 | (1.66, 2.50) | 1.23E-11 | 2.80 | (2.08, 3.78) | 1.07E-11 | 2.04 | (1.43, 2.92) | 8.13E-05 |
| TNFRSF1B | Tumor necrosis factor receptor superfamily member 1B | 2.50 | (1.91, 3.29) | 4.29E-11 | 2.00 | (1.68, 2.38) | 1.41E-14 | 2.59 | (1.80, 3.73) | 2.57E-07 |
| EFEMP1 | EGF-containing fibulin-like extracellular matrix protein 1 | 2.88 | (2.07, 4.02) | 3.37E-10 | 1.94 | (1.32, 2.85) | 7.49E-04 | 2.04 | (1.27, 3.27) | 3.01E-03 |
| B2M | Beta-2-microglobulin | 2.36 | (1.80, 3.09) | 3.89E-10 | 2.21 | (1.59, 3.09) | 3.14E-06 | 2.67 | (1.75, 4.07) | 5.10E-06 |
| TNFRSF1A | Tumor necrosis factor receptor superfamily member 1A | 2.24 | (1.74, 2.88) | 4.28E-10 | 2.48 | (1.76, 3.48) | 1.72E-07 | 2.25 | (1.51, 3.36) | 6.45E-05 |
| SVEP1 | Sushi, von Willebrand factor type A, EGF and pentraxin domain-containing protein 1 | 2.04 | (1.62, 2.56) | 1.30E-09 | 1.82 | (1.51, 2.20) | 4.94E-10 | 1.61 | (1.20, 2.15) | 1.33E-03 |
| MMP7 | Matrilysin | 1.68 | (1.42, 2.00) | 2.34E-09 | 1.52 | (1.23, 1.89) | 1.38E-04 | 1.24 | (1.05, 1.48) | 1.25E-02 |
| TREM1 | Triggering receptor expressed on myeloid cells 1 | 1.87 | (1.52, 2.29) | 2.36E-09 | 1.63 | (1.27, 2.09) | 1.33E-04 | 1.57 | (1.14, 2.16) | 6.07E-03 |
| SVEP1 | Sushi, von Willebrand factor type A, EGF and pentraxin domain-containing protein 1 | 1.93 | (1.54, 2.41) | 9.27E-09 | 1.94 | (1.58, 2.39) | 2.14E-10 | 1.49 | (1.12, 1.98) | 5.76E-03 |
| IL15RA | Interleukin-15 receptor subunit alpha | 2.03 | (1.59, 2.60) | 1.78E-08 | 1.99 | (1.50, 2.64) | 2.21E-06 | 1.60 | (1.15, 2.22) | 5.43E-03 |
| HAVCR2 | Hepatitis A virus cellular receptor 2 | 1.91 | (1.52, 2.39) | 1.88E-08 | 1.72 | (1.31, 2.25) | 9.55E-05 | 1.65 | (1.18, 2.32) | 3.61E-03 |
| TNFRSF1B | Tumor necrosis factor receptor superfamily member 1B | 1.88 | (1.50, 2.36) | 5.06E-08 | 1.98 | (1.64, 2.40) | 1.06E-12 | 2.69 | (1.87, 3.87) | 9.47E-08 |
| CNTN1 | Contactin-1 | 0.35 | (0.24, 0.51) | 7.30E-08 | 0.52 | (0.34, 0.81) | 3.27E-03 | 0.49 | (0.30, 0.78) | 2.68E-03 |
| HDGF | Hepatoma-derived growth factor | 1.70 | (1.40, 2.07) | 8.52E-08 | 1.48 | (1.12, 1.96) | 5.42E-03 | 1.61 | (1.19, 2.17) | 1.95E-03 |
| PSIP1 | PC4 and SFRS1-interacting protein | 1.74 | (1.42, 2.13) | 1.10E-07 | 1.61 | (1.21, 2.14) | 1.12E-03 | 1.54 | (1.16, 2.04) | 3.00E-03 |
| NBL1 | Neuroblastoma suppressor of tumorigenicity 1 | 1.79 | (1.44, 2.22) | 1.34E-07 | 1.69 | (1.36, 2.11) | 2.21E-06 | 1.76 | (1.34, 2.30) | 4.43E-05 |
| TMPO | Lamina-associated polypeptide 2, isoforms beta/gamma | 1.52 | (1.29, 1.78) | 2.95E-07 | 1.59 | (1.34, 1.90) | 1.43E-07 | 1.66 | (1.25, 2.19) | 3.92E-04 |
| SLAMF1 | Signaling lymphocytic activation molecule | 1.57 | (1.32, 1.87) | 2.99E-07 | 1.56 | (1.27, 1.92) | 3.16E-05 | 1.61 | (1.26, 2.06) | 1.36E-04 |

### Pathway analysis

The pathway analysis revealed 18 significantly enriched pathways, including Liver X Receptor/Retinoid X Receptor (LXR/RXR) Activation, Pathogen induced Cytokine Storm Signaling Pathway, Wound Healing Signaling Pathway, Acute Phase Response Signaling, Immunogenic Cell Death Signaling Pathway, IL-6 Signaling, peroxisome proliferator-activated receptor (PPAR) Signaling, and Crosstalk between Dendric Cells and Natural Killer Cells (**Figure 3 and Table S8.A**). For most pathways, activation was associated with a higher risk of sepsis (colored red in **Figure 3**). On the other hand, LXR/RXR Activation and PPAR Signaling are pathways relevant to anti-inflammatory signaling, and their inhibition was associated with a higher risk of sepsis (colored blue in **Figure 3**). When we used the 90 proteins that were replicated in the CHS cohort for pathway analysis, 5 pathways were enriched with |z-score| of ≥2 and 4 of them overlapped with the above 18 pathways. However, none of the 5 pathways remained significant after multiple testing correction (**Table S8.B**)

**Figure 3:**
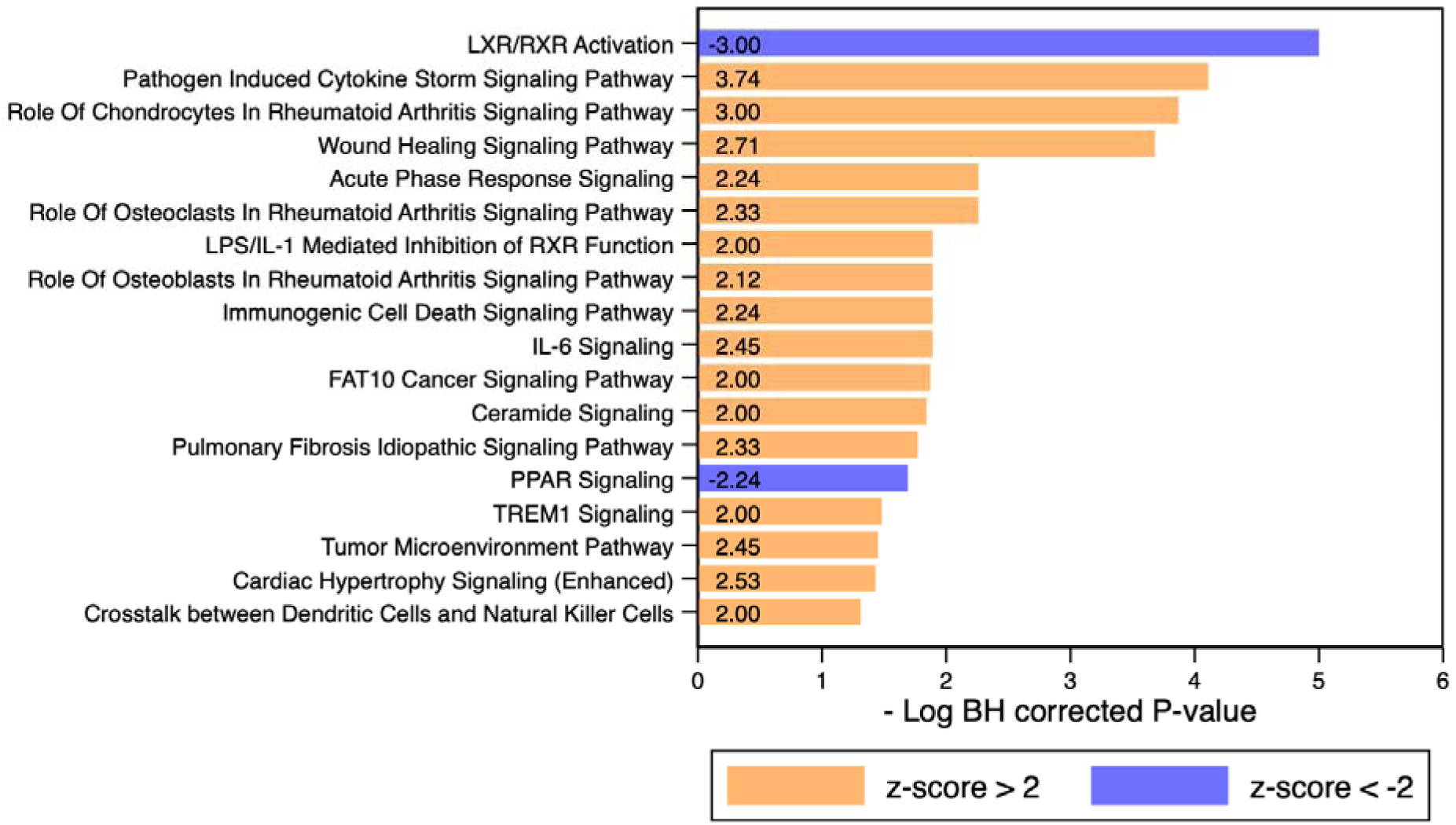
Enriched canonical pathways associated with risk of sepsis determined by the Ingenuity pathway analysis. Input parameters were comprised of the 175 proteins that were significant at ARIC visit 3 and visit 5. Among the 175 proteins, 2 were not mapped and 3 were duplicate identifiers, resulting in 170 proteins that mapped to the IPA database. A small p-value indicates that there are more proteins included in the pathway than expected by chance. Z-score quantifies the consistency in the directionality of an association (e.g., inhibition vs. activation) between observed and reference dataset. For example, a high positive z-score indicates that known activating proteins in a pathway consistently showed positive associations in the observed dataset. Threshold for statistical significance was a P value of <0.05 after Benjamini–Hochberg FDR adjustment. The threshold for Z-value was absolute z-scores of ≥2.

### Causal inference analysis

Of the 175 proteins significant both at visit 3 and visit 5, PWAS models were available for 102 proteins (i.e., 102 proteins had valid instruments and were linked to the protein levels)^19^ (**Table S9**). In these 102 proteins, the heritability of the gene (i.e., proportion of variation in protein levels explained by pQTLs) was generally low (median [IQI], 4.9% [2.0% and 11.0%]). Protein-wide association analysis did not discover proteins that were causally associated with the risk of sepsis based on FDR-corrected P value of <0.05, but highlighted 8 proteins at a nominal significance threshold of <0.05: CAPG, NCAM1, RNASET2, CTSZ, TNFRSF1B, AHSG, FBLN5, and IGFBP2 (**Table 2**).

**Table 2:** Protein-wide association study for risk of sepsis. Of the 102 proteins assessed in this analysis, only highlighted 8 proteins at a nominal significance threshold of <0.05 are presented in this table. P-values for the rest of 94 proteins are shown in Table S9.

| Gene symbol | Chromosome | SNPs (n) | Heritability of the gene (%) | R<sup>2</sup> for prediction model | PWAS z-score | PWAS P-value |
| --- | --- | --- | --- | --- | --- | --- |
| CAPG | 2 | 55 | 7.88% | 0.16 | -2.9323 | 0.00337 |
| NCAM1 | 11 | 115 | 4.84% | 0.13 | 2.89506 | 0.00379 |
| TNFRSF1B | 1 | 50 | 3.47% | 0.049 | -2.46657 | 0.0136 |
| AHSG | 3 | 78 | 11.00% | 0.19 | -2.4245 | 0.0153 |
| RNASET2 | 6 | 66 | 9.82% | 0.16 | -2.349926 | 0.018777 |
| FBLN5 | 14 | 75 | 0.94% | 0.022 | 2.09694 | 0.036 |
| IGFBP2 | 2 | 34 | 0.44% | 0.011 | -2.0968 | 0.03601 |
| CTSZ | 20 | 27 | 1.99% | 0.031 | 2.054 | 0.03997 |

### Predictive performance of protein models

Finally, we developed prediction models for risk of sepsis. For the base model, the c-statistic was 0.710 (**Table 3 and Table S10**). We then ran a LASSO regression model and identified 105 predictive proteins from the 669 proteins that were significant in the ARIC visit 3 cohort. When these proteins were incorporated into the model, the c-statistic rose to 0.795 (95% CI, 0.773 to 0.817), which was a significant improvement compared to the base model (Δc-statistic, 0.085 [95%CI, 0.068 to 0.101]).

**Table 3:** C-statistics for predicting risk of sepsis Change in the discover cohort (ARIC visit 3) and internal/temporal (ARIC visit 5) and external (CHS) replication cohorts. LASSO regression models selected the list of predictive proteins and their corresponding β coefficients, where the number of predictive proteins was determined by a tuning parameter λ. To balance between the predictive performance and model overfitting, we chose a λ that minimized the cross-validated partial likelihood deviance based on 10-folds cross validation. Approach 1 used the same set of predictors and regression coefficients from the discovery cohort for the validation cohorts. Approach 2 used the same set of predictors but allowed refitting regression coefficients within each validation cohort. The models were adjusted for age, sex, race, education attainment, body mass index, smoking status, eGFR, diabetes, hypertension, prevalent coronary heart disease, prevalent stroke, and prevalent heart failure.

| Approach/Models | ARIC visit 3 (Discovery) |  | ARIC visit 5 (Internal/temporal validation) |  | CHS (External validation) |  |
| --- | --- | --- | --- | --- | --- | --- |
| | C statistics | $\Delta C$ | C-statistics | $\Delta C$ | C-statistics | $\Delta C$ |
| **Approach 1** |  |  |  |  |  |  |
| Base model | 0.710 | Ref | 0.649 | Ref | 0.633 | Ref |
| LASSO model | 0.795 | 0.084 (0.068, 0.101) | 0.691 | 0.041 (0.025, 0.058) | 0.653 | 0.020 (-0.006, 0.046) |
| **Approach 2** |  |  |  |  |  |  |
| Base model | - | - | 0.661 | Ref | 0.652 | Ref |
| LASSO model | - | - | 0.754 | 0.093 (0.065, 0.121) | 0.732 | 0.080 (0.052, 0.108) |

This LASSO model significantly improved the c-statistics in the temporal validation analysis using ARIC visit 5 data (Δc-statistic, 0.041 [95%CI, 0.025 to 0.058]). This model improved c-statistic in the external validation cohort, but the improvement was not statistically significant (0.020 [-0.006 to 0.046]). However, the same set of 105 proteins considerably improved c-statistic when we refit the models (Δc-statistic, 0.093 [0.065, 0.121] from 0.661 to 0.754 for the ARIC visit 5 data; and 0.080 [0.052 to 0.108] from 0.652 to 0.732 for the CHS data) (**Table 3**).

For ARIC visit 3 data, we estimated the 10-year risk stratified by deciles of the predicted risk of sepsis. In the LASSO model, participants in the top decile of predicted risk had a 10-year sepsis risk of 5.4% whereas the risk was <0.5% in the first to fifth deciles (**Figure S4A**). These findings were also consistent in Cox proportional hazard analyses: the HRs comparing the bottom vs. top decile were 13.9 (95%CI, 8.0 to 24.3) for the base model, and 51.4 (95%CI, 24.1 to 109.7) for the LASSO model (**Figure 4A** and **Figure 4B**). These findings were consistent when using ARIC visit 5 data, although the risk gradients were less evident reflecting a higher baseline risk in this population (**Figure S5A and S5B**). For CHS replication cohort, the LASSO model showed good calibrations in the low predicted risk groups (i.e., first to sixth deciles) but not in the high predicted risk groups (i.e., seventh to tenth deciles). (**Figure S5C and S5D**).

**Figure 4:**
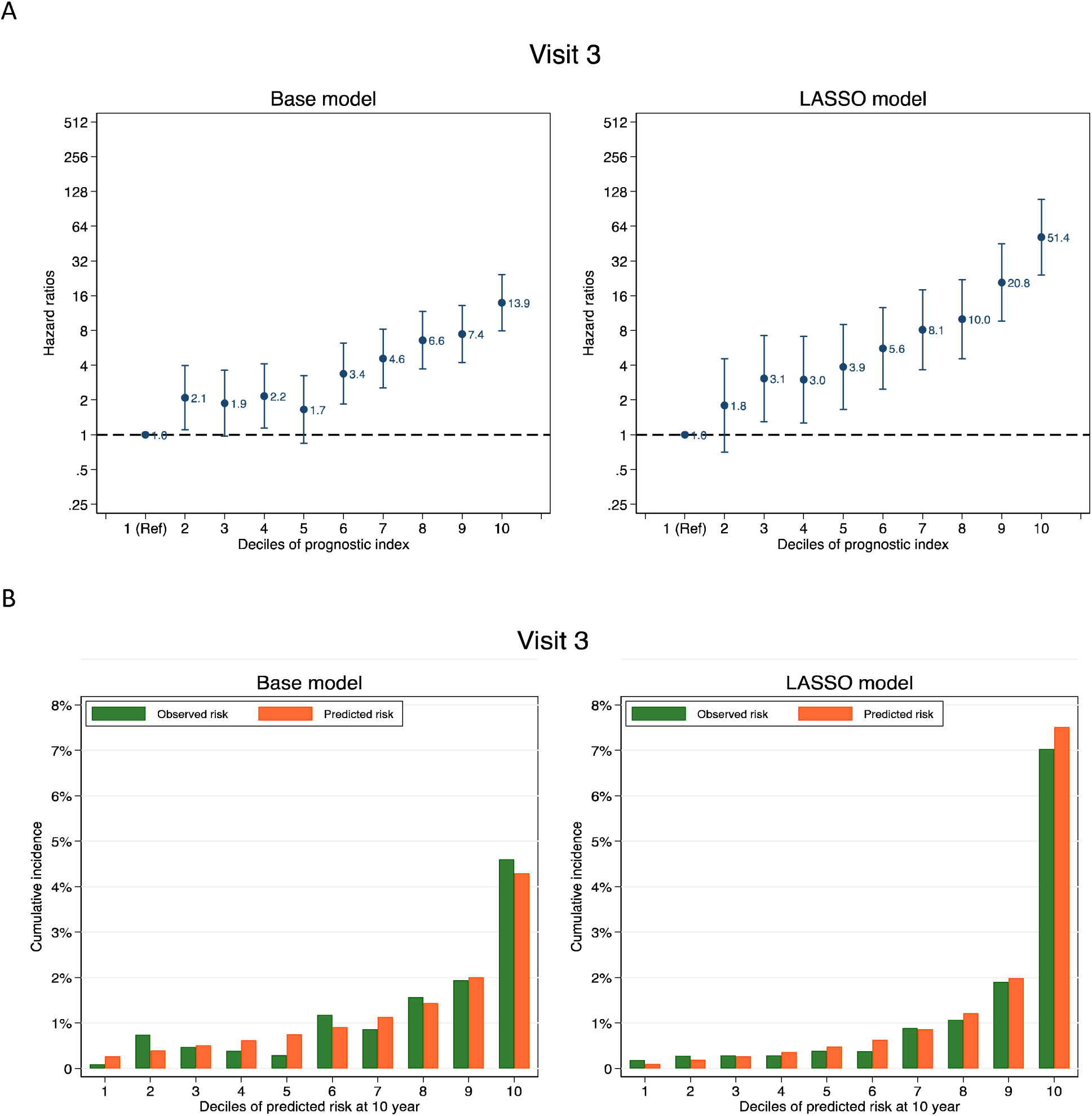
Adjusted hazard ratios and calibration of the base vs. LASSO prediction model: ARIC Study 1993-2019. The models were adjusted for age, sex, race, education attainment, body mass index, smoking status, eGFR, diabetes, hypertension, prevalent coronary heart disease, prevalent stroke, and prevalent heart failure. Calibration plots show the mean observed and predicted risk of sepsis within the discover cohort (ARIC visit 3).

## Discussion

In this study, we identified 669 proteins significantly associated with the risk of sepsis in midlife (visit 3) ARIC cohort. Of those, 175 proteins were replicated in late-life (visit 5) ARIC cohort, of which 90 proteins were replicated in an external late-life cohort of CHS. The top protein hits included 18 proteins and were involved in acute inflammatory processes in innate immunity.

Pathway analysis confirmed the significant enrichment of pathways relevant to the pro-inflammatory response, such as Cytokine Storm Signaling Pathway. Additionally, pathways related to the anti-inflammatory response, such as LXR-RXR activation, were also enriched for the risk of sepsis. Finally, we demonstrated that a prediction model incorporating proteomics data can discriminate individuals at risk of sepsis, particularly those with the highest risk (e.g., a 10-year risk of >5%).

Of the top 20 proteins (including 18 unique proteins), 16 proteins were positively associated with the risk of sepsis: for example, higher levels of TNFRSF1A and TNFRSF1B were consistently associated with a higher risk of sepsis. Although TNFR is a membrane-bound protein, the extramembrane portion of TNFRSF1A and TNFRSF1B is cleaved by the metalloprotease TNFα converting enzyme (TACE) to form soluble TNFRSF1A and TNFRSF1B. In an event of sepsis, the level of soluble TNFRSF1A and TNFRSF1B is markedly elevated in response to TNFα production.^40^ Soluble TNFR has capacity to bind TNFα and inhibit the excessive activation of acute TNFα signaling.^41^ Meanwhile, persistent TNF signaling activation can disrupt the translocation of NF-κB to the nucleus,^42,43^ and cause hypo-responsiveness to toll-like receptor signaling.^44^ Similar mechanisms of persistent stimulation leading to receptor desensitization have also been reported for chemokines,^45^ interleukins,^46,47^ and growth factors.^48^ Our findings support a hypothesis where chronic “irritation” of inflammatory pathway while free of the disease, as manifested by higher levels of inflammatory markers in the blood, may impair a healthy immune response against infection, thereby increasing susceptibility to sepsis.

On the other hand, some proteins such as EGFR and CNTN1 showed an inverse association with the risk of sepsis (i.e., higher levels were associated with a lower risk of sepsis). EGFR has been extensively researched as an oncogenic protein highly expressed in cancer cells, and it also plays a significant role in innate immunity. In healthy tissues, EGFR exhibits minimal expression, but its levels rapidly escalate through toll-like receptor signaling activation, triggering various responses such as interleukin activation, neutrophil recruitment, and epithelial repair.^49^ CNTN1 has been initially identified as a neuronal protein that regulates signaling between myelin and axon, and recently garnered attention as a potent oncogenic protein.^50^ Regarding its role in immunity, CNTN1 suppresses RIG-I and MAVS signaling in innate immunity,^51^ and the loss of *Cntn1* function resulted in global immune deficiency.^52^ However, both EGFR and CNTN1 are membrane-bound, and the mechanisms governing their release into the bloodstream, as well as the biological functions of their soluble forms, remain poorly understood.

Pathway analyses provide further insight into a pathophysiological landscape beyond the individual roles of proteins. Enriched pathways were consistently related to acute inflammatory pathways particularly in the innate immune response, such as LXR-RXR activation, cytokine storm signaling pathway, wound healing signaling pathway, and IL-6 signaling pathway. All of these pathways included at least one of the IL-1, TNFα, or IL-6, which are known to activate classic proinflammatory transcription factors such as NF-κB,^53^ Janus kinase/signal transducer and activator of transcription (JAK/STAT),^54^ PI3K/Akt and the mammalian target of rapamycin (mTOR).^55^

Interestingly, some enriched pathways, such as LXR-RXR activation and PPAR signaling, showed their inhibition to be associated with the risk of sepsis, rather than activation. Recent evidence has revealed that these pathways activate anti-inflammatory signaling by suppressing proinflammatory nuclear receptors, such as NFkB and other pro-inflammatory cytokines.^56^ Further, both LXR and PPAR are nuclear receptors (i.e., regulate gene transcriptions) that play central roles in lipid and glucose metabolism.^57^ For example, LXR is activated by cholesterol overload in macrophages, and induces the efflux of cholesterol and the expression/secretion of cholesterol transporters (e.g., apolipoprotein E, high-density lipoprotein [HDL] cholesterol).^57^ Epidemiological studies have demonstrated associations of dyslipidemia, such as low HDL, with the risk of pneumonia.^58^ Further, several medications that modulate lipid metabolism, such as statin^59^ and metformin,^60^ have been explored for the prevention or treatment for infections, although clinical trial data are limited.

The PWAS did not discover significant causal proteins for sepsis once accounting for multiple comparison. However, low heritability (<10%) of assessed proteins may limit a statistical power. Furthermore, the development of sepsis is influenced by non-host factors (e.g., pathogen factors). Nonetheless, highlighted proteins (i.e., unadjusted P-values < 0.05) including TNFRSF1B, are reasonably linked to immune response. For example, RNASET2 (Ribonuclease T2)^61^ and CTSZ (cathepsin Z, lysosome protease)^62,63^ are enzymes that degrade microbial proteins or RNAs to be recognized by toll-like receptors. CAPG,^64^ FBLN5,^65^ and NCAM1^66^ are primarily expressed in macrophages and natural killer cells, regulating cell motility, adhesion, and migration during initial immune response against pathogens. On the other hand, AHSG^67^ and IGFBP2^68^ are inhibitors of the insulin receptor or components of the insulin-like growth receptor, respectively.

These proteins have pleiotropic effects on cell growth signaling and lipid and glucose metabolism, but also modulate the immune response.^69^

Finally, we found that a prediction model based on proteins showed an ability to discriminate individuals at risk of sepsis, particularly those with the highest risk of sepsis: participants in the highest decile of predicted risk had a 10-year sepsis risk of 5.4%, whereas those in the low (e.g., first to fifth) deciles had a very low risk of sepsis (e.g., <0.5%). The model was not necessarily well-replicated in external older-aged cohort. Further, it should be noted an exploratory nature of this analysis since an immediate application of the prediction model may be limited since proteomics data are yet to be utilized in routine clinical care or health screening settings.

Nonetheless, the same set of 105 proteins considerably improved c-statistics, including the external CHS cohort, when we refit the models, which is noteworthy given that the risk for communicable diseases should be determined by both host and pathogen factors.

Several limitations should be acknowledged. First, the observational studies are subject to residual confounding and measurement error. Second, the generalizability of our findings may be restricted to White and Black individuals residing in the US community. Third, our outcome ascertainment relied on ICD codes recorded in the primary diagnostic position on discharge records. This approach may offer high specificity but low sensitivity. Strengths of this study include our ability to relate blood protein measurements to perform an extensive analysis combining proteomics analysis, pathway analysis, and protein-wide association analysis, and develop a prediction model incorporating proteomics data for sepsis risk.

In conclusion, in this large-scale proteomics analysis, levels of acute inflammatory proteins measured during routine visits were associated with the subsequent incidence of sepsis. An increased risk of sepsis associated with the inhibition of anti-inflammatory pathways, such as LXR/RXR Activation warrants further mechanistic investigation.

## Data Availability

Pre-existing data access policies for each of the parent cohort studies specify that research data requests can be submitted to each steering committee; these will be promptly reviewed for confidentiality or intellectual property restrictions and will not unreasonably be refused. Please refer to the data sharing policies of these studies. Individual level patient or protein data may further be restricted by consent, confidentiality or privacy laws/considerations. These policies apply to both clinical and proteomic data.

## Acknowledgements

The **Atherosclerosis Risk in Communities Study** has been funded in whole or in part with Federal funds from the National Heart, Lung, and Blood Institute, National Institutes of Health, Department of Health and Human Services, under Contract nos. (75N92022D00001, 75N92022D00002, 75N92022D00003, 75N92022D00004, 75N92022D00005). SomaLogic Inc. conducted the SomaScan assays in exchange for use of ARIC data. This work was supported in part by NIH/NHLBI grant R01 HL134320. The authors thank the staff and participants of the ARIC study for their important contributions. This **Cardiovascular Health Study** research was supported by NHLBI contracts HHSN268201200036C, HHSN268200800007C, HHSN268201800001C, N01HC55222, N01HC85079, N01HC85080, N01HC85081, N01HC85082, N01HC85083, N01HC85086, 75N92021D00006; and NHLBI grants U01HL080295, R01HL087652, R01HL105756, R01HL103612, R01HL120393, U01HL130114, HL144483 with additional contribution from the National Institute of Neurological Disorders and Stroke (NINDS). Additional support was provided through R01AG023629 from the National Institute on Aging (NIA). A full list of principal CHS investigators and institutions can be found at CHS-NHLBI.org.

The content is solely the responsibility of the authors and does not necessarily represent the official views of the National Institutes of Health.

